# A general model for the demographic signatures of the transition from pandemic emergence to endemicity

**DOI:** 10.1101/2021.07.06.21260071

**Authors:** Ruiyun Li, C. Jessica E. Metcalf, Nils Chr. Stenseth, Ottar N. Bjørnstad

## Abstract

Anticipating the medium- and long-term trajectory of pathogen emergence has acquired new urgency given the ongoing COVID-19 pandemic. For many human pathogens the burden of disease depends on age and prior exposure. Understanding the intersection between human population demography and transmission dynamics is therefore critical. Here, we develop a realistic age-structured (RAS) mathematical model that integrates demography, social mixing and immunity to establish the suite of possible scenarios of future age-incidence and burden of mortality. With respect to COVID-19, we identify a plausible transition in the age-structure of risks once the disease reaches seasonal endemism, whether assuming long-lasting or brief protective immunity, and across a range of assumptions of relative severity of primary *versus* subsequent reinfections. We train the model using diverse real-world demographies and age-structured social mixing patterns to bound expectations for changing age-incidence and disease burden. The mathematical framework is flexible and can help tailoring mitigation strategies countries worldwide with varying demographies and social mixing patterns.

**One Sentence Summary:** A shift of COVID-19 risks to younger age-classes in future endemic circulation.

## Introduction

The unfolding pandemic caused by SARS-CoV-2 threatens to be one of the biggest challenges of our time. Mounting evidence suggests a seemingly inevitable resurgence of disease towards endemism in the foreseeable future (*1,2*). Identifying the age and burden profiles that may define the years ahead could help improving response preparedness, both for this pandemic, and for future emerging pathogens.

A fundamental signature of COVID-19, the disease associated with SARS-CoV-2, is the age manifestation of the burden of infection and morbidity. Following infection by SARS-CoV-2, there is a clear signature of increasingly severe outcomes and fatality with age (*3-5*). Historical emergence of acute respiratory infections indicates that age-incidence patterns during virgin epidemics can be very different from endemic circulation (*6,7*). This motivates efforts to bound the potential future age-circulation and fatality to understand the evolving health burden.

Predicting age-circulation in the near and mid future (e.g., 1-5 years since emergence) requires realistic age-structured (RAS) mathematical models that includes characterization of immunity following (re-)infection. Empirical evidence from seasonal coronaviruses indicates that prior exposure may only confer short-term immunity to reinfection, allowing recurrent outbreaks (*8,9*). Despite this, prior exposure may prime the immune system to provide protection against severe disease (*8,10,11*), and thus possibly reduce the public health burden of future recurrences. Here, we propose an age-structured multi-compartmental SIRS model that allows for projections for future age-circulation and disease burden of SARS-CoV-2 virus under various plausible scenarios. We frame our model around a balance between simplicity and flexibility and structure our analysis accordingly. First, to develop a base-line for the transition of age-dependent risk over long-term dynamics, we explore outcomes for a ‘rectangular demography’ (where survival is complete until a maximum age, resulting in a rectangular age pyramid and constant population size) and ‘homogeneous mixing’ (where individuals have equal probability of contact with individuals of all other ages). The purpose of this mass-action, homogeneous mixing model is to provide a baseline for thinking about transitions in age-incidence over time. As per all mass-action models of virgin epidemics, the initial post-epidemic trough is unrealistically deep. Using a power-scaling law to allow for spatial and social clustering as suggested by Liu et al. (*12*) will obviously alleviate this, but add parameters that doesn’t add to the overall take-home message. The subsequent step is to incorporate demographic and social complexities, including realistic age pyramids and assortative contacts among age groups, both derived from country-specific data.

Our general model projects age-incidence and thus age-morbidity patterns into the future according to chains of differential equations:

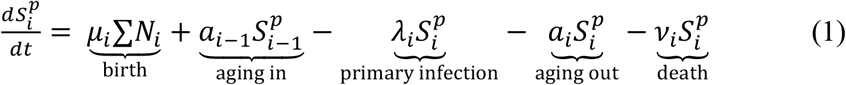

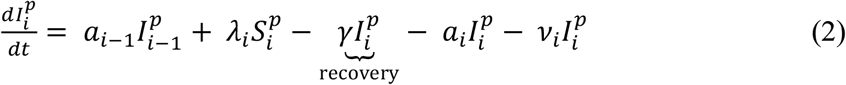

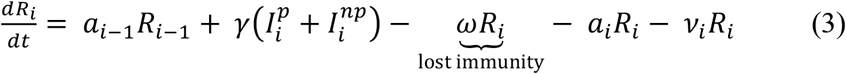

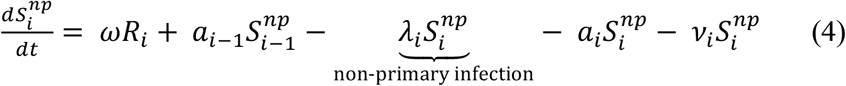

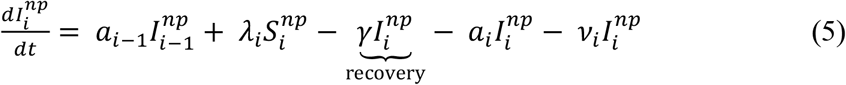

where 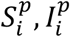 are the number of susceptible individuals and primary infections in age group *i*. The recovered individuals (*R*_*i*_) may lose immunity and return to susceptibility 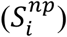 after an average protected period of 1/*ω* and subsequently be liable to reinfection; accordingly, 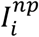 is the number of non-primary infections in age group *i*. The force-of-infection on susceptibles in age-class which is the rate at which any susceptible of age *i* will be infected is 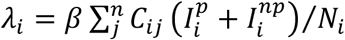, where *β* is the baseline rate of transmission given by *β* = *R*_0_*γ* and *C*_*ij*_ is the normalized contact rate between age group *i* and *j*. In the below illustration we assume an 80-year life expectancy and thus a birth rate *μ*_*i*_ = 1/80 year^-1^ at which people are born to the youngest group in a population of size *N*_*i*_ (i.e *μ*_*i*_ is 0 for all *i* > 1). For the baseline model, we assume *a*_*i*_ to be the age-specific rate of aging with a 1-year duration, *v*_*i*_ is a rate of natural mortality (we assume *v*_*i*_ = 0 for all *i* until the rectangular age end-point), and 1/*γ* to be the average duration of infection (taken to be 7 days in this analysis). To map our model to realistic demographies and social mixing patterns, we parametrize the model based on a broad range of countries. Details of model parameters are provided in Table 1.

**Table 1.**
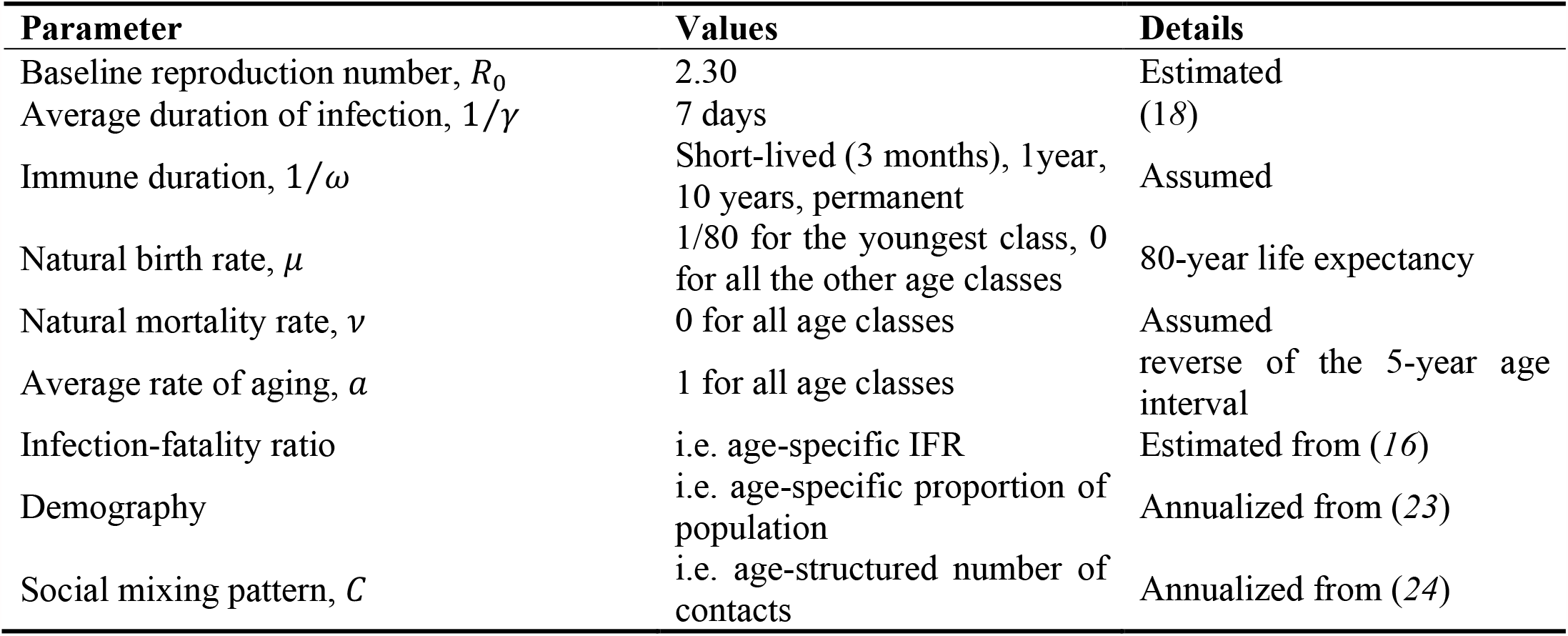
SIRS Model parameters.

| Parameter | Values | Details |
| --- | --- | --- |
| Baseline reproduction number, $R_0$ | 2.30 | Estimated |
| Average duration of infection, $1/\gamma$ | 7 days | (18) |
| Immune duration, $1/\omega$ | Short-lived (3 months), 1 year, 10 years, permanent | Assumed |
| Natural birth rate, $\mu$ | 1/80 for the youngest class, 0 for all the other age classes | 80-year life expectancy |
| Natural mortality rate, $\nu$ | 0 for all age classes | Assumed |
| Average rate of aging, $a$ | 1 for all age classes | reverse of the 5-year age interval |
| Infection-fatality ratio | i.e. age-specific IFR | Estimated from (16) |
| Demography | i.e. age-specific proportion of population | Annualized from (23) |
| Social mixing pattern, $C$ | i.e. age-structured number of contacts | Annualized from (24) |

## Results

We first identify the broad consequences of the intersection of immunity and burden over immediate, medium and longer terms (year 1, 10 and 20 years, respectively), explicitly considering immune scenarios that differ in the degree to which immunity prevents reinfections and/or attenuates severe cases, and then consider realistic demographics and social mixing for 11 different countries chosen to span diverse demographies. Across a 20-year horizon, we assess age-specific risk during a virgin epidemic, medium-term and a scenario of long-term endemic circulation.

Prevalence is predicted to surge during a virgin epidemic but then recede in a diminishing wave pattern as the spread of the infection unfolds over time towards the (probably seasonally-varying) endemic equilibrium (Fig. 1A-B). Depending on immunity and demography, the virgin epidemic the RAS model predicts a strikingly different age-structure than the eventual endemic situation (Fig. 1C-D; Fig. S1). When considering overall disease burden in the population during a probable transition from emergence to endemicity, our model highlights the importance of three main axes of variability/uncertainty: immune duration, demography and social mixing. Over the course of emergence, the shift in the age-profile of risk of infection and disease is largely dependent on the extent of infection-blocking and disease-reducing immunity. During the transition to endemism in a scenario of long-lasting immunity (assumed permanent or 10-year), the young – who for SARS-CoV-2 suffer a mild burden of disease – is predicted to have the highest rates of infection once the disease dynamics moves towards the steady-state (Fig. 1C; Fig. S1A), as older individuals are protected from infection by prior infection. If immunity to reinfection is brief (assumed short-lived 3-month or 1-year), changes in disease severity due to prior exposure is the main driver of changes to age-structured risk and long-term burden of mortality. The possibility of rapid reinfection, and severe outcomes on reinfection would heighten long-term circulation and continued high-risk infection among adults, though could modulate the age profile of risk over time (Fig. 1C; Fig. S1A). In contrast, if disease symptoms on reinfection are attenuated, the burden of disease may decay over time even if duration of sterilizing immunity is short-lived and reinfection is frequent. In the latter scenario the age-profile of primary infections will define the shifting risk over time, primary infection recedes to younger individuals as an emerging acute respiratory infection moves towards endemicity (Fig. 1D; Fig. S1B).

**Fig. 1.**
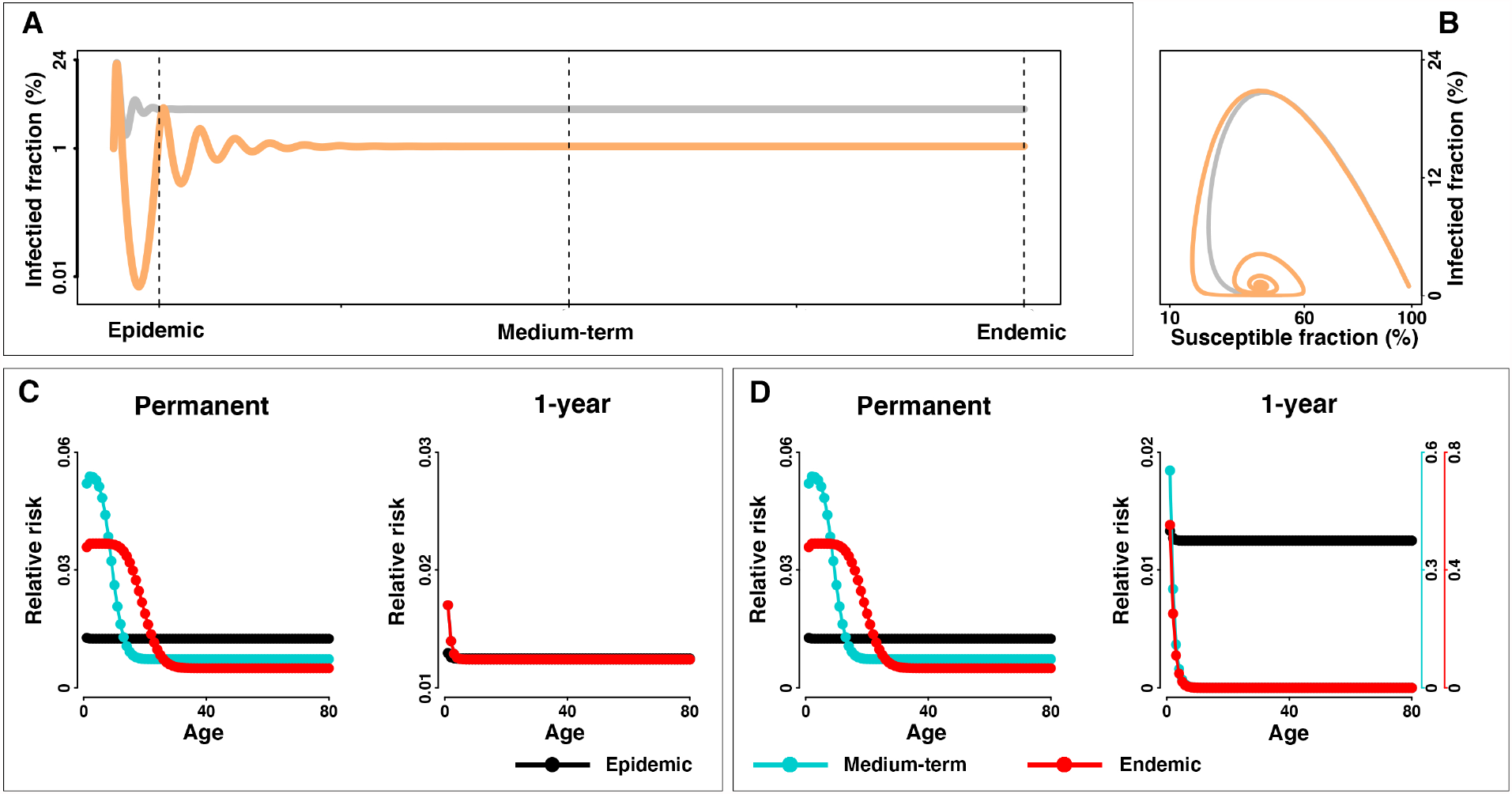
Trajectory of infected fraction and transitions in age-structure of the risk from virgin epidemic to endemic equilibrium. **(A)** Infected fraction for an outbreak is simulated with *R*_0_ = 2.3, 1/*γ* = 7 days, and a short-lived (i.e. 3-month) (grey) and 1-year (orange) immunity duration over 20 years. The SIRS model is parameterized with rectangular demographic structure and homogeneous social mixing pattern. Dashed lines indicate different stages of disease dynamics. For visualization, only trajectories in scenarios of short-lasting (i.e. short-lived and 1-year) are presented. **(B)** shows the infected versus susceptible fraction. If primary and non-primary infections have similar illness, **(C)** shows relative risk (i.e. age-specific infected fraction relative to the population-wide fraction) among age groups in the virgin epidemic, medium-term and endemic stage under scenario of permanent and 1-year immunity duration, respectively. If non-primary infections are less severe, **(D)** shows relative risk from primary infections. Relative risk among age groups under scenario of permanent, 10-year, 1-year and short-lived immunity duration are shown in the Supplement Material (see Fig. S1).

Our general model framework allows for robust predictions regarding transition in age-profile of risk in the face of either short/long-term protective immunity, reduction of severity of disease given previous exposure, and consideration of the range of countries with their different demographies and social mixing patterns (Fig. 2, Fig. S2-S6). Broadly speaking we find that immune scenarios are the dominant driver of transitions in age-dependence and risk towards endemicity, though our framework to incorporate realistic demographies and social mixing patterns to modulate the relative risk is likely to provide a critical infrastructure for policy decision making.

**Fig. 2.**
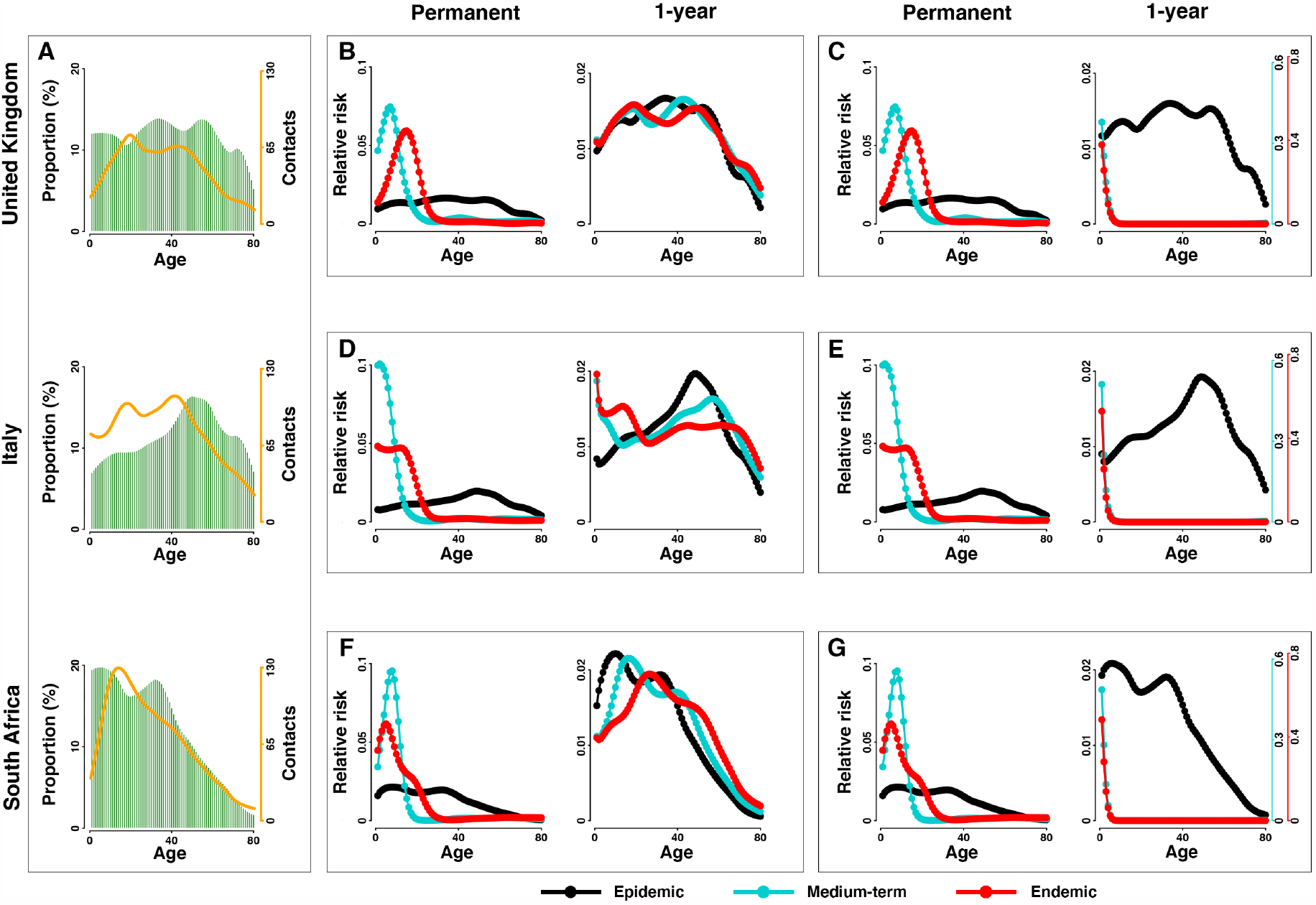
Transitions in age-structure of the risk in different countries. With **(A)** demographic structure (greens bars) and social mixing pattern (orange lines) in United Kingdom, Italy and South Africa, (**B-G)** show the relative risk among age groups in the virgin epidemic, medium-term and endemic stage in scenario of permanent and 1-year immunity duration. Risk from **(B, D, F)** all infections and **(C, E, G)** only primary infections are explicitly distinguished. Relative risk among age groups under scenario of permanent, 10-year, 1-year and short-lived immunity duration are shown in the Supplement Material (see Fig. S2).

Assuming prior exposure reduces severity of respiratory reinfections, the model’s projected transition is broadly consistent with those documented in several historical respiratory pandemics. In particular, the critical importance of both age and prior exposure on disease burden and mortality following the 1918 pandemic has been well-characterized: the elderly were protected by immunity from previous exposure to an earlier A/H1N1 related strain but within some years the overall burden of mortality receded (*6,13,14*). Ongoing genomic work following on (*15*) tantalizes that the million-killing1889/90 pandemic could have been caused by the emergence of HCoV-OC43 which is now an endemic mild repeat-infecting coronavirus.

For SARS-CoV-2 preparedness our model provides a robust framework for scenario analyses for the future. Irrespective of parametric uncertainties, the burden of mortality will peak during the virgin epidemic period (Fig. 3-4). The predicted magnitude of this peak is moderately affected across a plausible range of immune durations and immunity-modulated severity upon re-exposure (Fig. S7-S8). By contrast, post-pandemic burden during endemicity is shown to be strongly dependent on immune-function and prior infection history as it affects infection-probability and disease severity. Milder disease from reinfections (Fig. 3, Fig. S7) would give rise to decreasing mortality due to the reduction of severe cases, while burden of mortality over time may remain unchanging if primary infections do not prevent reinfections or mitigate severe disease among the elderly (Fig. 4, Fig. S8). In this bleakest scenario, excess deaths due to continual severe reinfections that results from the continuous replenishment of susceptibles via waning of immunity to reinfection will continue until effective pharmaceutical tools are available.

**Fig. 3.**
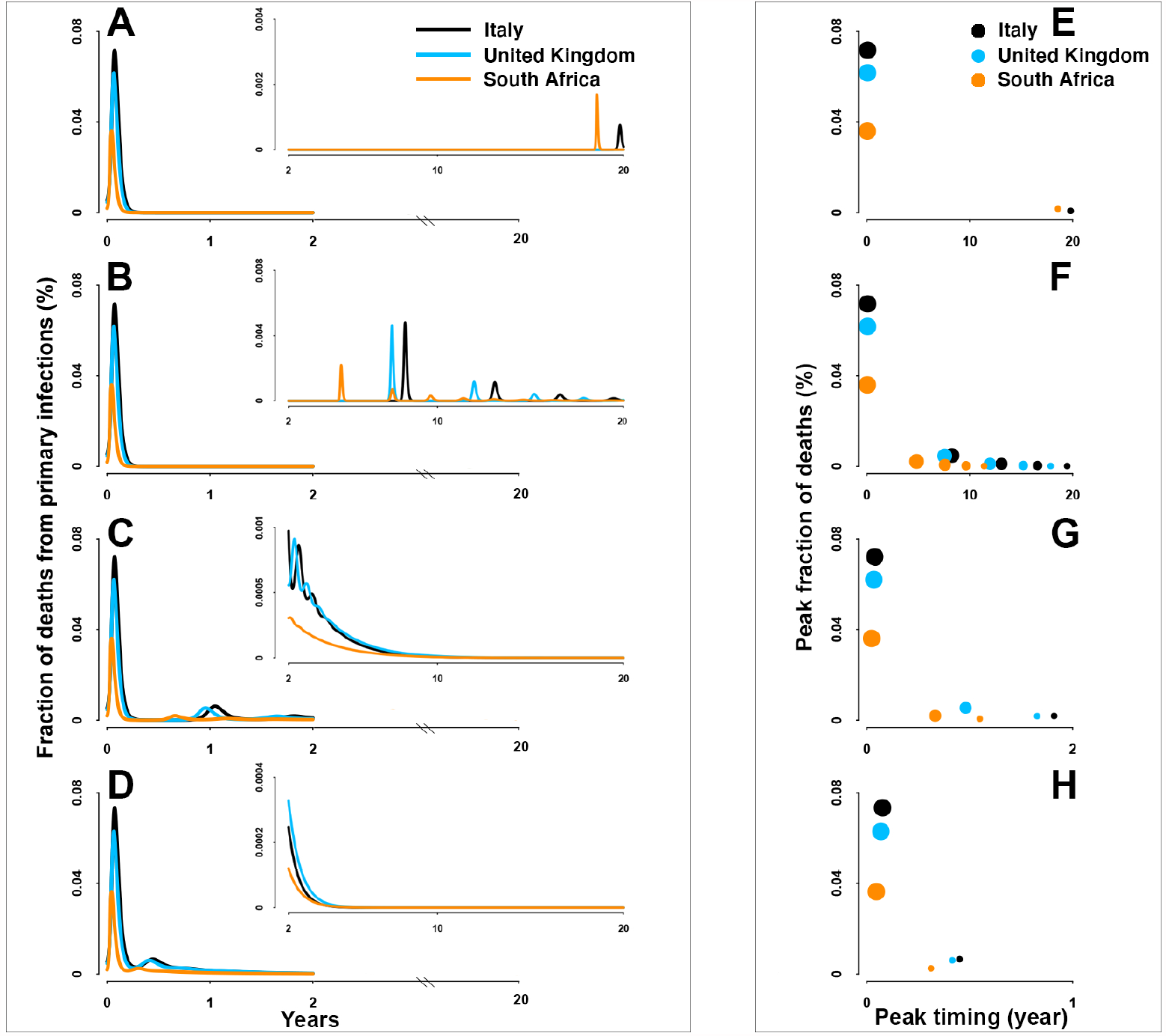
Fraction of deaths from primary infections. (**A-D)** Overall fraction of deaths over 20 years and (**E-F)** the timing and magnitude of consecutive peaks in scenario of permanent, 10-year, 1-year and short-lived (i.e. 3-month) immunity duration, respectively. Countries with different demographies and social mixing patterns are distinguished by colour: Italy (black), United Kingdom (blue) and South Africa (orange). For visualization, insets show trajectories following the first 2 years.

**Fig. 4.**
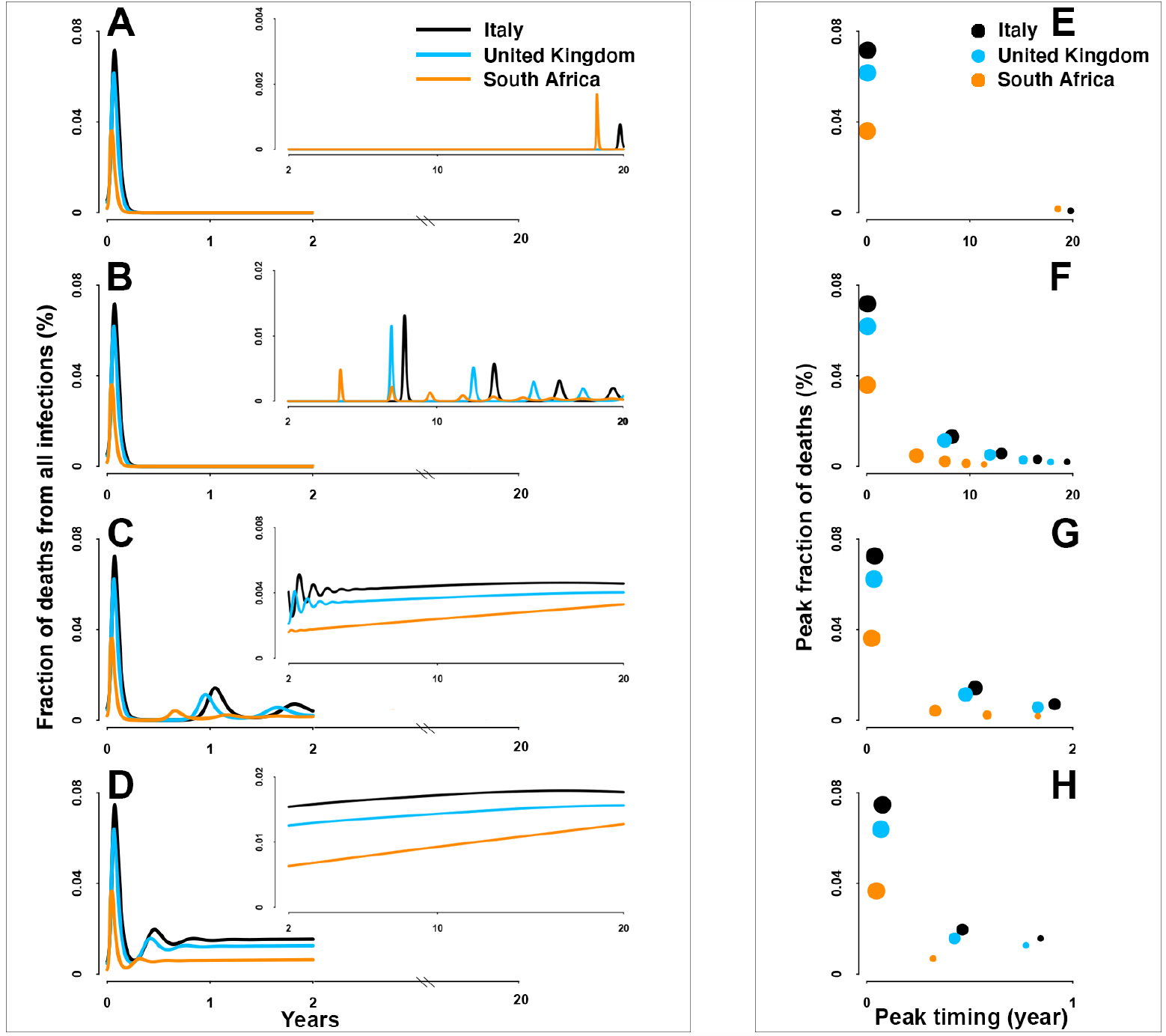
Fraction of deaths from all infections. Same as **Fig 3** but for deaths from both primary and non-primary infections.

A final insight from our detailed RAS model is that regardless of immunity and mixing, the population-level burden of mortality may differ among countries because of varying demographies. Given the marked increase of infection-fatality ratio (IFR) with age, countries with older population structure would be expected to have a typically larger fraction of deaths than those with relatively younger population structure (once corrected for differences in public health infrastructure). Consistent with this, South Africa, partially due to its younger population structure, has a lower fraction of deaths as compared with older populations such as Italy. These “deaths disparities” among demographies are largely invariant over the unfolding pandemic, though young people would be predicted to contribute most to burden in the endemic era. When comparing the relative importance on the overall burden of mortality, we show that the varying demographies is a key determinant of the disparities among countries (see Supplementary Text “*Relative effect of demographies and social mixing patterns*”; Fig. S9-S10).

## Discussion

Our realistic age-structured SIRS model provides a general framework to explore various scenarios for the possible unfolding of the current and future pandemic crises in the face of country-specific demographies, social mixing patterns and the inevitable main unknowns for any emergence such as immune duration and immune mitigated reduction in severity of disease. Through the integration of age structure, social mixing and immunity, our projections using SARS-CoV-2 as a focus for considering the broader issue, we highlight how risk will shift over time to different age-classes that may suffer different burden of disease during an endemic state. Such a shift will be very strong if immunity is long-lived but also of great public health significance if immunity to reinfection wanes, yet previous exposure attenuates severity of disease.

By highlighting a wide range of scenarios, our model framework is a robust scaffolding to help improve preparedness and mitigation of the current and future pandemics. Furthermore, our realistic age-structured SIRS model makes critical contributions to understanding how the highly variable social context, particular demography and age-structured mixing patterns, may modulate current and future disease burden. Building upon our framework (including the detailed and documented code provided in the supplement), health authorities will have a powerful and flexible tool to conceptualize future age-circulation, strengthening context-specific preparedness and deployment of interventions. The model will handily accommodate additional uncertainties/ variabilities as evidence is accumulated in the coming months and years.

Our RAS model makes several assumptions. First, we focus on the infection-blocking immunity. Incorporating realistic immunity efficacy with respect to susceptibility, transmission and severity is an important direction for expansion. Second, we assume a general formulation for the epidemic model. This should be considered as the starting point for the extension to encompass disease-specific mechanisms. Furthermore, we assume a homogeneous susceptibility to infection, clinical fraction and infection *vs* case-fatality ratio across age classes. Relaxing our assumptions by explicitly consider age-specific heterogeneities (*8, 16,17*) is an important future direction. Last, we assume an exponential decay of immunity within each age class which leads to a Gamma distributed loss over time. We believe this is a realistic model that still needs to be further refined with the mounting clinical and empirical studies. (see *Supplementary Text* for details of the assumptions and directions for further extensions).

## Materials and Methods

### Model parameterization

#### Transition rates

In the base model, we assume an 80-year life expectancy and thus a birth rate *μ*_*i*_ = 1/80 year^-1^ at which people are born to the youngest group in a population of size *N*_*i*_ (i.e *μ*_*i*_ is 0 for all *i* > 1). We assume *a*_*i*_ to be the age-specific rate of aging with a 1-year duration (i.e. *a*_*i*_ = 1 for all *i*). *v*_*i*_ is a rate of natural mortality which is assumed 0 for all age classes until the rectangular age end-point (i.e. *v*_*i*_ = 0 for all *i*). 1/*γ* is the average duration of infection which in the analysis taken to be 7 days (*18*). We further explore its variability on epidemiological trajectories (see Sensitivity analysis). In the RAS model with country-specific population pyramid and contacts over age, we retain the assumption of zero mortality across ages below the maximum age, the same birth rate to the youngest group and the same aging rate across countries, as this will result in a broadly consistent age structure (appropriate to human demography where transients play out extremely slowly).

#### Estimating the reproduction number

The reproduction number is a critical parameter for our model projections. Social distancing is well documented to impact transmissibility (*19*) and many countries implemented such interventions during the build-up of the virgin epidemic. Given this, we assume that the effective reproduction number i.e. the level of transmissibility, on day *t, R*_*t*_, is linked to the reduced mobility on that day, *m*_*t*_, via:

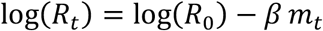

where *R*_0_ is the basic reproductive number in the absence of behavioural changes and *β* is the transmission rate. Reduced mobility leads to reductions in the effective reproduction number. When the reduction of mobility/mixing is 0%, *R*_0_ provides the baseline transmissibility parameter. We use China as the reference point.

We used daily confirmed cases in China (*20*) and extract mobility from the Baidu database (*21*) in the period of January 1^st^ – March 5^th^, 2020. With the cases we estimate *R*_*t*_ using a 14-day sliding time window using *EpiEstim* package (*22*), assuming a mean of 5.1 days and standard deviation of 5.3 days of the serial interval (*23*). We then exclude estimates of *R*_*t*_ prior to January 15^th^, 2020 in subsequent analysis, given the limited number of cases and thereby large uncertainty of *R*_*t*_. We also trim the estimates of *R*_*t*_ after February 20^th^, 2020 (i.e. four weeks after lockdown on January 23^rd^, 2020) when mobility rebounded but was not a strong corelate of reductions in *R*_*t*_ since then. With estimates of *R*_*t*_ and mobility data from January 15^th^ – February 20^th^, 2020 (Fig. S12), we establish the transmissibility-mobility association and estimate *R*_0_ using generalized linear model (GLM) with a negative binomial link function. Note that we do not tend to explicitly fit the documented cases nor optimize every transmission parameter; instead, we capture *R*_0_ to characterize the overall basic transmissibility. The estimated *R*_0_ is subsequently used as the baseline to simulate dynamics of COVID-19 in the age-structured SIRS model framework (*see below*). Furthermore, we examine how different demographies are predicted to modulate country-specific *R*_0_ ’s away from the early Chinese baseline (see Supplementary Text “*Variation of transmissibility among countries*”).

#### Demographics and age-structured social mixing patterns

To fully characterize the long-term age-circulation across the globe, we select 11 countries across a broad range of demographic and social mixing patterns. The countries cover Asia (China, Japan and South Korea), Europe (Spain, United Kingdom, France Germany and Italy), North America (United States), South America (Brazil) and Africa (South Africa). For these countries, we collected age pyramids from the statistics of the United Nations (*24*) and country/age-specific number of contacts from Prem et al. (*25*). We further annualized these data to generate the finer age profile into the 1-year age brackets necessary for model predictions (Fig. S13).

#### Age-specific infection-fatality ratio

Pilot studies have shown an increased infection-fatality ratio with age. We collect the posterior estimates of infection fatality ratio (IFR) from Verity et al. (*17*), and subsequently project them onto the 80 age groups in our study to predict burden.

### Model projections

Given the realistic age-structured SIRS equations defined by eqs (1)-(5), we numerically integrate the model to predict dynamics of COVID-19 for the next 20 years using a variety of scenarios spanning a range of current unknowns. For each scenario, simulation was initialized with 1% infections and 0.1% recovered individuals, i.e. *S*^*p*^ (0) = 0.989, *I*^*p*^ (0) = 0.01, *R* = 0.001, and *S*^*np*^ (0) = *I*^*np*^ (0) = 0.

For initial insights we studied the base model using a rectangular demography (i.e. in the absence of infection everybody is expected to live to the age of 80, resulting in a rectangular age pyramid and constant population size) and homogeneous mixing (i.e. individuals have equal probability of contact with individuals of all other ages) under four different durations of immunity. i.e. the short-lasting immunity assumed as (i) short-lived (3-month) or (ii) 1-year, and the long-lasting immunity assumed as (iii) 10-year or (iv) permanent (or life-long). Of note, in the scenarios where reinfection is possible, functional immunity to disease may still vary (*8,10,11*). Given this, we explicitly consider two scenarios that differ in the severity of reinfections. We firstly assume an independence of disease severity from prior exposure, so the burden of disease depends on the sum of both primary and non-primary infections. Alternatively, we assume prior immunity may mitigate disease severity. In which case, milder reinfections is assumed of no contribution to shaping the epidemiological trajectories, and thus public health burden depends on age-profiles of primary infections.

Next, we contextualize the transition in age-circulation for the 11 selected countries (*see* “*Demographics and age-structured social mixing patterns*”). We add greater demographic and social complexity to the base model, by initiating the population with country-specific age pyramids and social mixing patterns obtained as described above. We then simulate the models with a broadly consistent age structure where where transients play out extremely slowly, by retaining the assumption of zero mortality across ages below the maximum age and the same birth rate to the youngest group across countries (see “*Transition rates*”). This helps titrate how these variables may lead to varying patterns among countries. Relative risk among age groups is defined as the infected fraction in each age group relative to that in a population as a whole. To assess plausible transitions towards endemicity, we estimate relative risk in the first, tenth and 20^th^ year following emergence (hereafter as the virgin epidemic, medium-term and the probable endemic phase, respectively). Finally, we project the trajectories of deaths in the selected countries under a variety of immune scenarios. The population-level fraction of deaths, i.e. burden of mortality, is estimated by multiplying the age-specific infected fraction with IFR. We assumed an invariant IRF for primary and non-primary infections. Consistent with above assessment of changing age-structure, we examine the scenario with different duration of immunity and possible mitigation of illness due to prior exposure. To evaluate the relative importance of demography and social mixing pattern, we further simulate the model by using the assumed homogeneous mixing patterns (see Supplementary Text “*Relative effect of demographies and social mixing patterns*”).

### Sensitivity analyses

We validate our findings and insights by examining the uncertainty that may arise from the assumed duration of infection and model formulation. More specific, we investigate the dynamics of disease burden by assuming an array of the average duration of infection, including 5, 9, 11 days. Additionally, we formulate a SEIRS model by explicitly incorporating the exposure (E) component and asymptomatic infections:

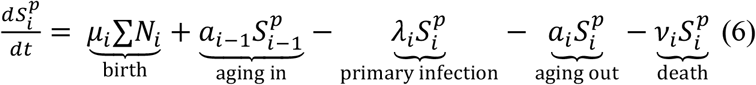

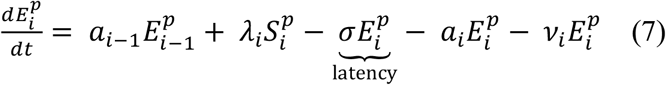

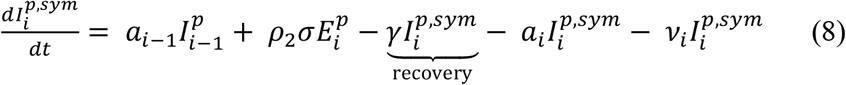

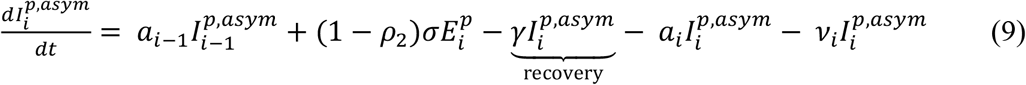

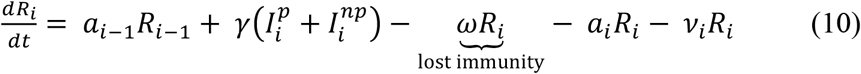

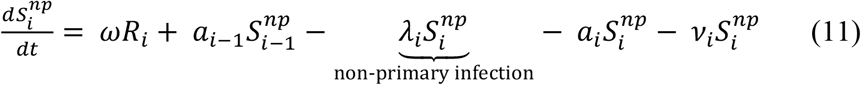

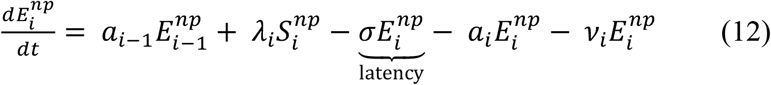

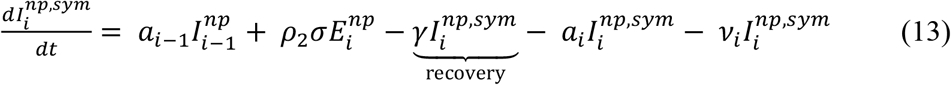

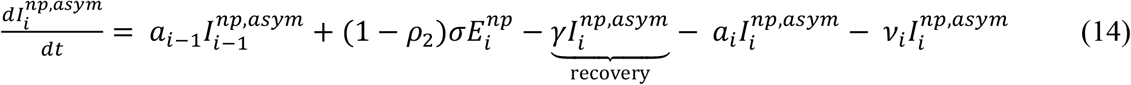

For simplicity, we assume that the average duration of incubation (*δ*) is 6.4 days (*26*), the infectiousness of asymptomatic (*ρ*_1_) is half (50%) of that of symptomatic infections, and the proportion of asymptomatic infections (*ρ*_2_) is 40% (*27*). The force of infection on susceptibles in age-class *i* is defined by 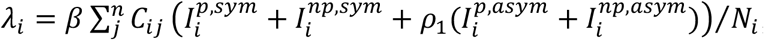, where superscript *sym* and *asym* denotes the symptomatic and asymptomatic infections, respectively. *β* is the baseline rate of transmission given by *β* = *R*_0_*γ* and *C*_*ij*_ is the normalized contact rate between age group *i* and *j*. We simulate the model and estimate the age-specific risk in the scenario of the rectangular demography and homogeneous mixing.

## Data Availability

Not applicable.

## Acknowledgments: Funding

This study was supported by grants from the Huck Institute of the Life Sciences, Penn State University through the Seed-funded COVID-19 Projects; Research Council of Norway through the COVID-19 Seasonality Project (Reference No. 312740). The funders of the study had no role in study design, data collection, data analysis, data interpretation or writing of the report.

## Author contributions

R.L. built the model, collected data, finalized the analysis, interpreted the findings and wrote the manuscript. C.J.E.M. built the model, interpreted the findings and wrote the manuscript. N.C.S. and O.N.B. designed the study, interpreted the findings, commented on, and revised drafts of the manuscript. All authors read and approved the final manuscript. The corresponding author had full access to all the data in the study and had final responsibility for the decision to submit for publication.

## Data and materials availability

The data and code that support the findings in this study has been made openly available at https://github.com/ruiyunli90/Corona-ages.

## Supplementary Materials

Supplementary Text

Figures S1-S13

References (*28*-*29*)

